# From Food Aid Invoices to Impact: A New Method to Quantify Caloric Intake from Community Kitchens in Conflict Affected Sudan

**DOI:** 10.1101/2025.06.10.25329322

**Authors:** Yamna Ouchtar, Ahmed Mohammed, Samantha Olson, Ruwan Ratnayake, Laura Skrip, Maysoon Dahab

## Abstract

In conflict-affected regions, especially contested and inaccessible settings, where data collection is often limited, monitoring food availability and identifying populations at high nutritional risk remains a significant challenge. In Sudan, where armed conflict by military actors persists into a third year, a complex humanitarian crisis has unfolded, marked by hindered international humanitarian access and a deficiency of up-to-date information about the ground reality. In this context, locally-led food aid interventions play a vital role in mitigating food insecurity, especially in besieged settings with poor or no access to external food aid. ‘Community Kitchens’ or *Takaya’s* as they are known locally are a cornerstone of these interventions, providing cooked meals neighbourhood by neighbourhood. We present a novel operational method that uses food purchase invoices and attendance records from these kitchens to estimate caloric intake among beneficiaries, offering an alternative means of estimating caloric intake where traditional nutrition information is not possible. By linking data from handwritten food aid invoices, ingredient lists, and attendance records, we calculated the average caloric intake of beneficiaries and tracked changes over time and across multiple kitchens. Our methodology demonstrates the feasibility of using invoice-based modelling to understand food aid distribution and calorie intake from existing operational data.

Our method represents a rapid and flexible way of measuring the impact and trend of food availabilities in otherwise inaccessible settings. Our results indicate that community kitchen food aid significantly and rapidly improved caloric intake among those who accessed it. However, overall kitchen coverage remains limited with variations on impact linked to funding restrictions, increases in food prices and attacks on kitchens and their staff. This work highlights the importance of community-led food aid and the utility of operational data where widescale nutritional data collection is not possible. It also reinforces the urgent need to sustain and increase community kitchen interventions, especially in response to escalating operational challenges, to the already nutritionally compromised populations they serve.

## Background

Sudan is experiencing one of the world’s most severe and complex humanitarian crises (1,2), driven by compounding and overlapping forces of armed conflict, economic collapse, and climate shocks. Longstanding nutritional vulnerability in Sudan reflects these historical, political, and socio-economic factors. Even before the war, in 2022, 49% of the population across the country were food insecure, with nearly 60% of households in the urban centre of Khartoum state struggling to meet their basic needs (3,4). As the war enters its third year, an estimated 25 million people, more than half of the country’s population, are thought to be food insecure (5).

Adult caloric intake, a key marker of nutritional vulnerability, has also been steadily decreasing. In 2014, the average daily caloric intake of adults in urban areas was 2,247 (6), declining steadily thereafter. The 2018 economic crisis accelerated this trend, with devaluation of the Sudanese currency, and a 150-200% rise in the price of staple foods such as wheat or millet making households in Khartoum and elsewhere struggle to meet their basic needs (7). After the war in Khartoum State, where the conflict remains volatile, 67% of smallholder farmers were unable to plant in the last growing season, and nearly half reported no intention to cultivate in the future (8).

The escalation of violence between the Sudanese Armed Forces (SAF) and the Rapid Support Forces (RSF) has intensified these vulnerabilities, by displacing 16.1 million people (9), disrupting livelihoods, destroying infrastructure and the already fragile health system (10,11). Meanwhile, humanitarian access is severely constrained, with bureaucratic obstruction and insecurity rendering many areas unreachable by international agencies (12).

An often-overlooked aspect of the crisis is the disruption of data collection and reporting, which has severely constrained humanitarian monitoring and response efforts. Established tools for ground-level assessment—such as the Standardised Monitoring and Assessment of Relief and Transitions (SMART) methodology (13)— have been significantly constrained by aforementioned challenges. These constraints have created critical blind spots, preventing the timely identification of nutritional needs in sensitive areas. The collapse of key famine early warning systems, notably FEWS NET, following the suspension of funding by the US Government, when access to reliable data was most urgently needed, has further weakened food security monitoring (14–16). This situation is exacerbated by the politicization of data: denial of famine conditions and obstruction of humanitarian access by parties to the conflict, as well as the recent withdrawal of the SAF from the Integrated Food Security Phase Classification (IPC) process, a standardized, consensus-based framework used to classify the severity and drivers of food insecurity, have exacerbated fears and encouraged reluctance to share data (17). The resulting information vacuum masks the scale of the nutritional crisis and prevents a scaled and needs-based response.

In the absence of functional state-led or internationally coordinated humanitarian mechanisms, local responses have become essential lifelines. Community kitchens (CKs), local interventions coordinated by volunteer networks and supported by diaspora-led fundraising, sometimes in collaboration with international partners (18), are the most important of these. Despite facing significant operational obstacles, including insecurity, communication breakdowns and chronic resource shortages, these kitchens continue to distribute hot, ready-to-eat meals in many parts of Sudan (19,20).

In this context of acute need and systemic data collapse, we aim to contribute to a new evidence base by analyzing operational data, namely food purchase invoices, from CK operators in one besieged setting in Khartoum. Our aim is to explore what these unconventional data sources can reveal about food access, caloric availability and local response capacity in a rapidly deteriorating nutritional landscape.

### Geographical and temporal scope

Secondary operational data was provided by Hadhreen a national Sudanese civil society organization working with a network of CKs operating across Sudan (21). Hadhreen has organised and supported CKs through a standard process (see Figure 1) in Sudan since November 2023 (22). Starting with a few kitchens, the initiative quickly expanded to over 200 kitchens in several states, serving diverse communities with support from several donors, each with specific reporting requirements. The kitchens operate through a decentralised, volunteer-led system and face significant operational constraints, including insecurity and limited connectivity (23).

**Figure 1:**
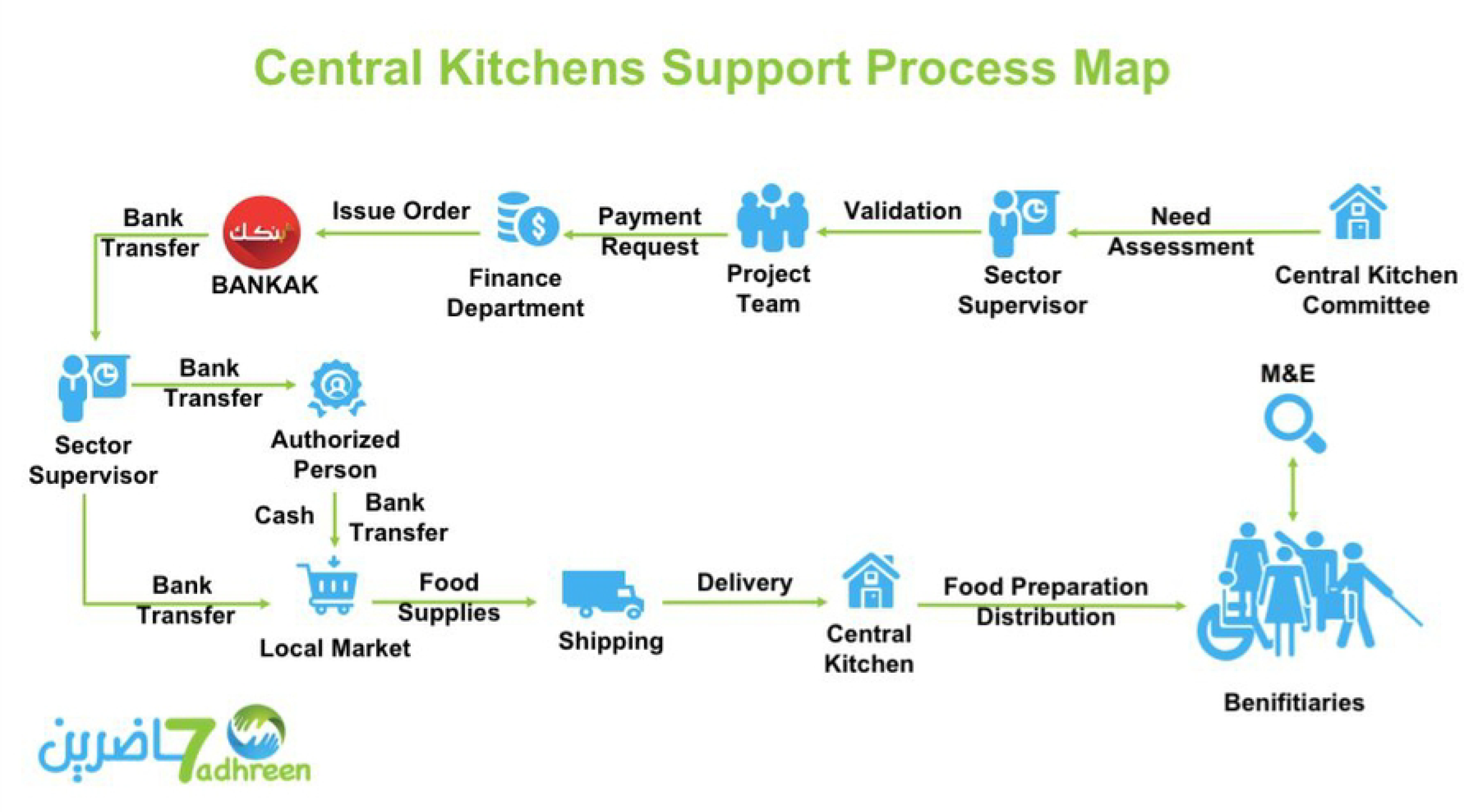
Hadhreen central community kitchen support process map (21).

For our analysis, we selected Hadhreen-supported CKs in the Jebel Awlia (JA) locality. This locality is one of the most food insecure areas of Khartoum State, heavily affected by active fighting and infrastructure damages bridge (24–26), leaving the population entrapped and heavy reliant on emergency food assistance through CKs. Assessments conducted by Hadhreen between October and November 2024 showed that CKs were the main source of food for beneficiaries, due to the absence of alternative food sources and the unaffordability of staple items in local markets (27). This aligns with IPC reports released in December 2024, classifying parts of Jebel Awlia (Mayo and Alingaz) to be ‘at risk of famine’ (28). Despite the growing need, outside assistance has been severely restricted with just two food aid conveys arranged by WFP reaching the area since the start of the war in April 2023 (29,30). The vulnerability of households in Jebel Awlia predates the war. Growing economic instability after 2018, meant that by 2020, 88% of households reported incomes insufficient to meet basic needs, with over 80% spending more than 65% of their income on food, highlighting the extreme vulnerability and chronic food insecurity in the area, now worsened by conflict and economic collapse (31). Considering these historical vulnerability, alongside current context and survey data, we considered Jebel Awlia to be a relatively closed food system. In addition, Jebel Awlia stands out for having a large number of active CKs. These factors it a particularly suitable setting for analysing the nutritional impact of CKs over time.

## Materials and Methods

To estimate food aid and caloric intake during the ongoing conflict in Sudan, this study uses an approach based on procurement invoices from CKs in a locality besieged by fighting and under control of the RSF in Khartoum State. Our methodology consists of five essential elements: 1. geographical and temporal scope; 2. acquisition and pre-processing of data from kitchen invoices; 3. estimation of food quantities and caloric content; 4. estimation of beneficiary population; and 5. estimation of pre-war caloric intake for contextual comparison.

### Data Processing

Our analysis was based on a detailed review of purchase invoices from Hadhreen- supported CKs operating in Jebel Awlia. To address operational challenges, Hadhreen uses a flexible digital workflow to securely manage data and ensure continued access for team coordinators and partners. Kitchen supervisors use WhatsApp to communicate and manage procurement, sharing daily updates, purchase orders and invoice information with Hadhreen through voice notes, photos and text messages. Hadhreen digitalizes photos of purchase invoices and purchased items, as well as counts of the number of meals distributed daily in each kitchen. These are stored in a Cloud folder using a unique code identifier number for each kitchen, which is used consistently across all operations. The code is based on the International Organization for Standardization (ISO) 3166-2 codes (32) and uses the standard state and locality abbreviations (33) followed by a unique kitchen number. In addition, to meet donor requirements, each food item was standardised with unique names and weight units, then converted to metric tonnes to allow aggregation and reporting by locality.

Building upon this operational workflow, we then developed a semi-automated data pipeline to process purchase invoices into data. First, we automatically translated and transcribed invoice details into structured purchase records using open-source code (Marian-MT transformer) from Hugging Face (34). Second, we converted the purchase records into machine-readable formats. Third and finally, we used Python scripts to extract and compile variables on types and quantity of food items into Excel spreadsheets for reporting and analysis. The digitisation process also involved cleaning and compiling daily meal counts into usable format.

A summary of the data collection and processing workflow is illustrated in Figure 2.

**Figure 2.**
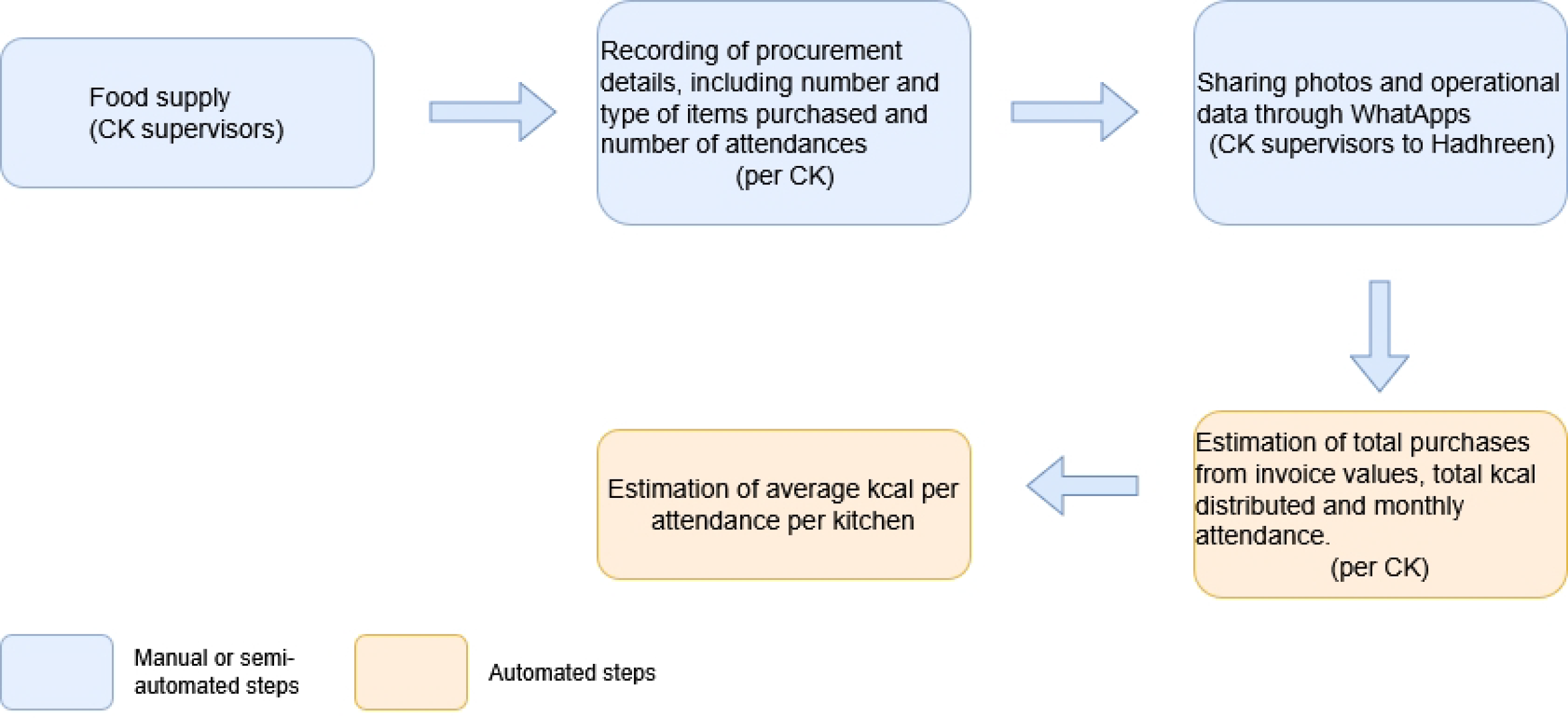
Summary of the pipeline for data collection, processing, and analysis.

### Estimation of food quantities and caloric content

To estimate the caloric value of the food distributed by CKs and convert individual food items into their caloric equivalents, we used NutVal 4.0, a software package developed by United Nations High Commissioner for Refugees (UNHCR) and the WFP for emergency nutrition planning (35), alongside food composition tables for Sudan (36). For each item listed in the kitchen invoices, the calorie values were calculated in kilocalories (kcal). These values were then aggregated across all invoices to generate the total calorie availability per kitchen and per supply cycle.

To understand the variation in food availability over time, we calculated the approximate intervals between consecutive supply invoices. This allowed us to estimate the frequency of replenishment and, by extension, the distribution of calories over time, a necessary step in tracking calorie trends throughout the study period. Thus, the model incorporates a key assumption concerning the rate of consumption of the food distributed. Based on field observations and key informant accounts from kitchen supervisors, we assumed a typical consumption window of 7 to 14 days between deliveries. This assumption provides a fundamental basis for estimating daily calorie availability per beneficiary.

### Estimation of beneficiary population

Initial estimates of the number of beneficiaries were drawn from Hadhreen’s 2024 baseline registration in Jebel Awlia. These mean values of the number of beneficiaries per CK were obtained by registering the household heads or family members and then applying simple linear interpolation method to estimate family sizes, resulting in a household size of five members. Attendance records from two subsequent distributions were used to refine and validate this household size estimate.

To deal with missing attendance data in some CK, we applied interpolation and imputation techniques, using a backward and forward fill approach on a per-kitchen basis (37). In addition, we used population estimates from the WorldPop dataset, scaled to projections for 2024 (38) to estimate the total population of Jebel Awlia and calculate the proportion potentially reached by CK interventions relative to this local population.

### Estimation of pre-war caloric intake for contextual comparison

To assess the impact of the conflict on food access, we estimated pre-conflict average calorie intake using national urban reference values from 2014 (2,247 kcal/day for urban adults) (6) and 2017 (1,893 kcal/day) (7). Due to lack of continuous surveillance, we applied linear interpolation between time points, adjusting for food inflation based on historical price indices (sorghum and millet). We validated this approach using an earlier 2009 nutrition survey that provided baseline estimates for food consumption trends.

In addition, to contextualize and cross-validate our estimates, we integrated conflict data from the Armed Conflict Location & Event Data Project (ACLED) dataset (39), accounting for the number violent events. Additionally, we analysed food price data obtained from WFP reports, normalizing and converting prices to USD (based on 2009 values) to compare with prices derived from invoices of the Hadhreen community kitchens and associated funding levels.

Given the inability to quantify food availability from sources other than CKs, we developed multiple scenarios to represent beneficiary caloric intake under different assumptions. These scenarios estimate the proportion of pre-war caloric availability accessible to beneficiaries, assuming they receive 25%, 50%, or 100% of pre-conflict levels.

## Ethics

Ethics approval was provided by the London School of Hygiene and Tropical Medicine (LSHTM) Ethics Committee for the use of public and non-public secondary data (ref. 31426). All analyses in this paper were done in R (40) and Python (41). All data and analysis scripts, are available from the authors: https://github.com/yamnao/food_extraction_sudan.

## Results

Our analysis (see Figure 3) indicates a significant decline in average daily calorie intake in Sudan’s urban areas since the baseline in 2014. This downward trend accelerates considerably after 2022, correlating with a sharp rise in staple food prices.

**Figure 3.**
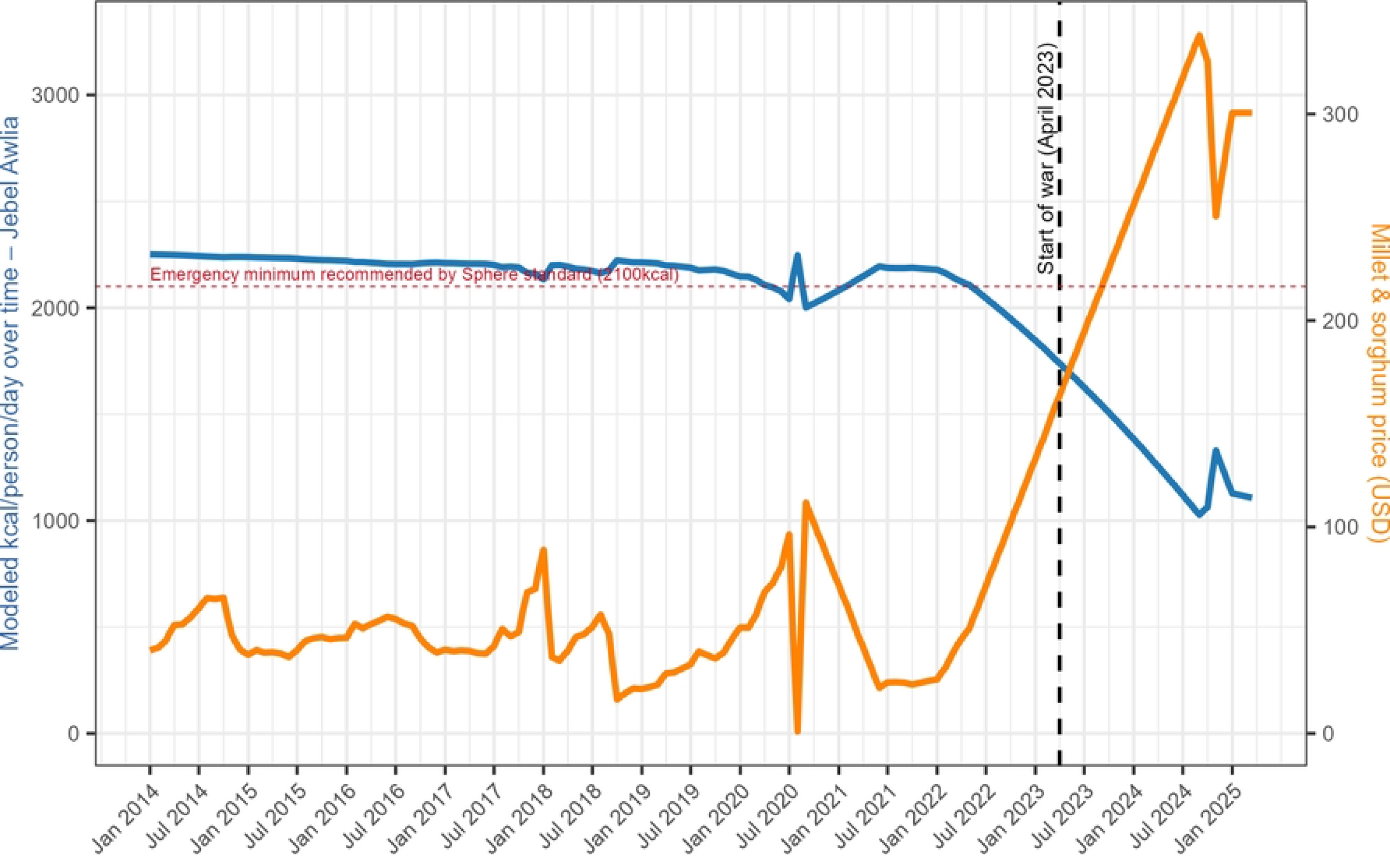
Modelling of kcal intake per person per day using linear interpolation between 2014 and 2017 kcal references scaled by USD price of millet and sorghum.

The estimation of caloric availability through CKs was informed by data from 318 purchase invoice and attendance records from 57 Hadhreen supported kitchens in Jebel Awlia between August 2024 to March 2025. It was estimated that CKs supplied between a high of 1,250 kilocalories per person per day (kcal/person/day) in September 2024 and a low of 500 kcal/person/day in December 2024 (Figure 4). Some kitchens reached distributions exceeding 1,400 kcal/person/day which corresponded with the periods of highest levels of funding availability. However, these levels varied considerably across both time and geographic location, driven by disparities in available funds and operational disruptions, including insecurity.

**Figure 4.**
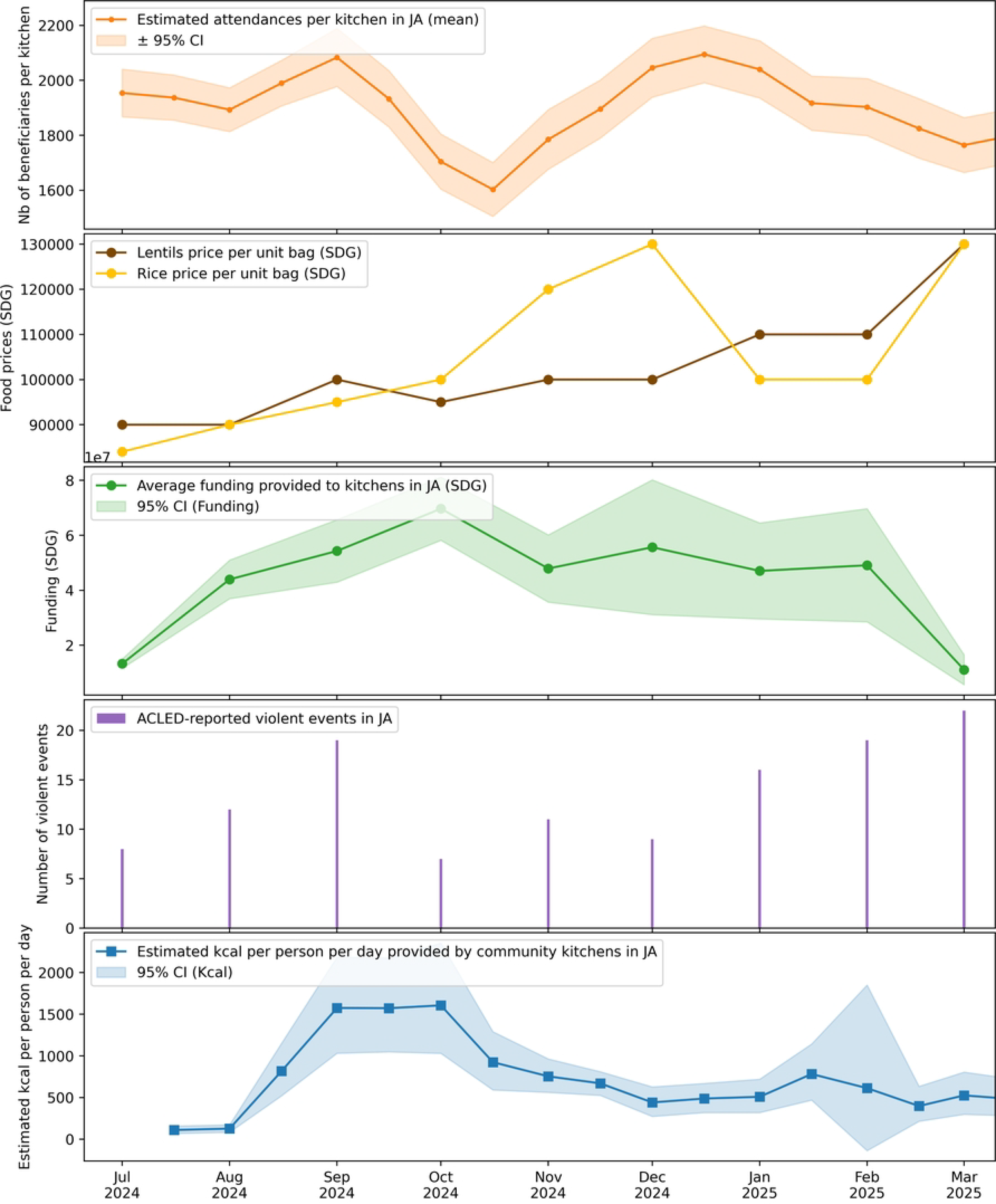
Estimated kcal per person per day provided by community kitchens, compared to attendance, food prices, funding levels, and frequency of violent attacks on kitchen reported by ACLED (39).

Despite limited dietary diversity - typically restricted to five main ingredients (rice, lentils, oil, onion and spices), nutritional analysis revealed that the protein and fat content of food provided was generally adequate in relation to emergency nutritional standards (Figure 5). However, while the overall macronutrient profile aligns with recommended values, the low per-person quantity of food intake suggests that individual daily requirements are unlikely to be met.

**Figure 5.**
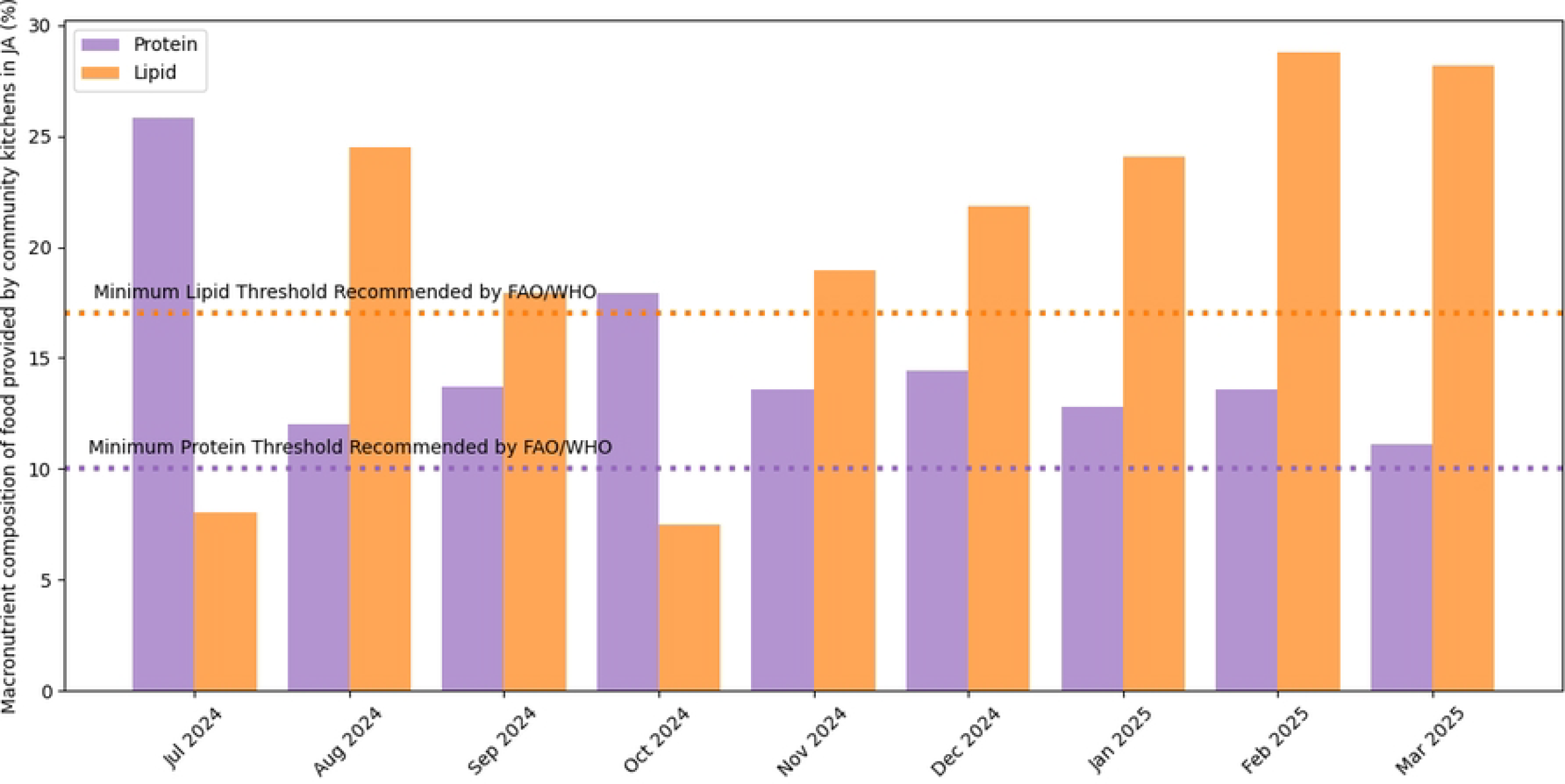
Macronutrient composition of food provided by community kitchens in Jebel Awlia and comparisons with FAO/WHO recommendations (42).

To better understand the possible scenarios, we integrated estimates of food availability from other sources (i.e., access to 25%, 50%, or 100% of prewar caloric intake), in addition to community kitchen food distribution, as shown in Figure 6.

**Figure 6.**
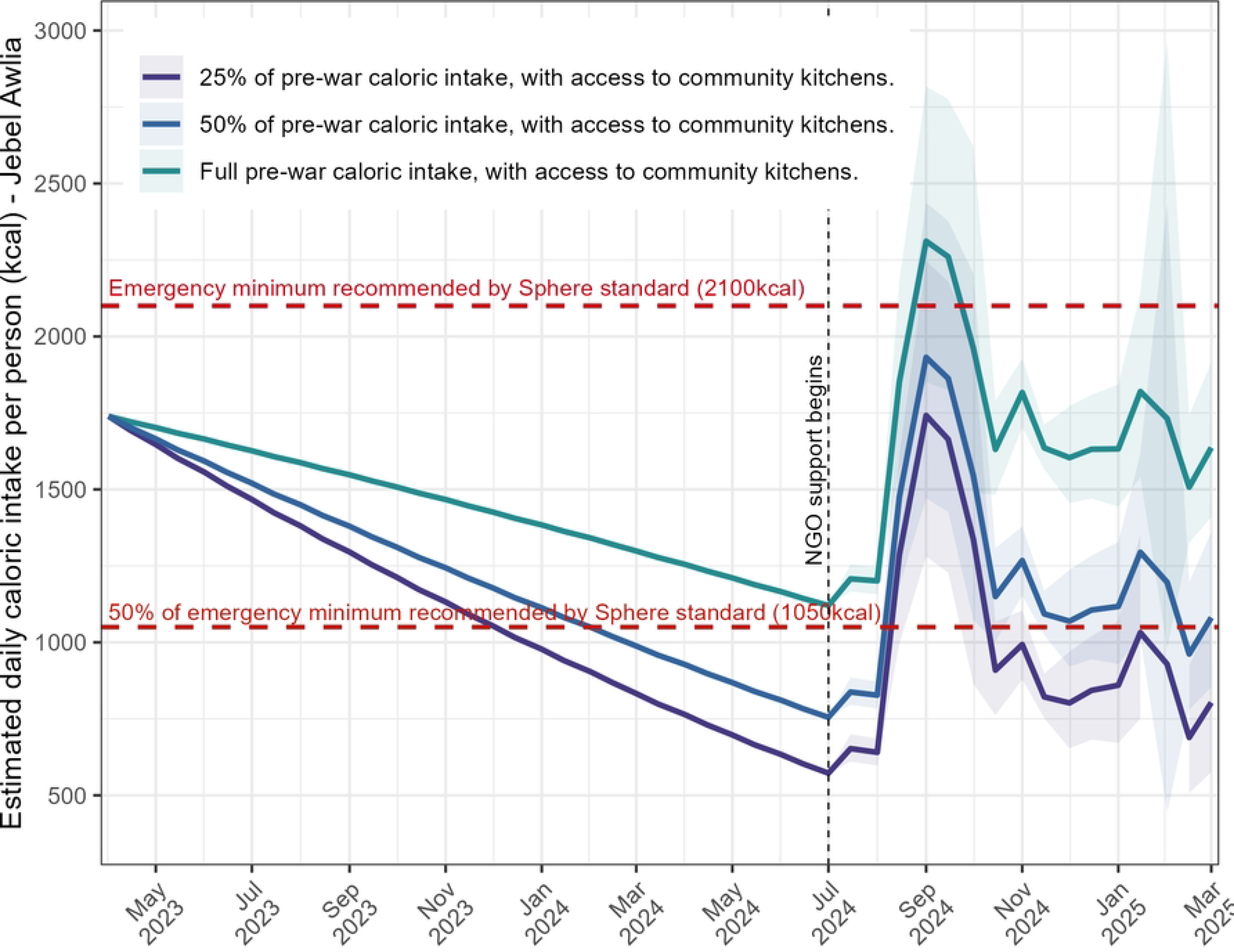
Estimation of kcal per person per day during the war-period in Jebel Awlia, in Sudan, using the kcal intake modelled by linear interpolation scaled by USD price of millet and sorghum and kcal intake provided by community kitchens.

## Discussion

Our analysis suggests that community kitchens provided critical additional kcal and thus played a vital role in stabilising access to food for thousands of people over the period studied. In many cases, they helped to maintain pre-war levels of daily calorie intake, particularly in the early months analysed. Early reports of community kitchen starting in Jebel Awlia appeared on social media in December 2023 (43,44), indicating that some caloric supplementation likely occurred prior to our observed period. Without these kitchens, most households would likely have faced extreme calorie deficits much earlier in the conflict.

In terms of outside relief, there are very few humanitarian actors providing assistance in Jebel Awlia. Due to the long-entrapped nature of the area (24), only two World Food Programme (WFP) convoys with food assistance are known to have reached Jebel Awlia; one in in August 2023 (45) and another in December 2024 (46). The later convey of 28 UN trucks carrying approximately 637 metric tons of food aid and additional medical supplies, marked the first significant delivery since the start of the conflict. This aid was intended to reach about 78,000 people in the Mayo and Alingaz districts (47,48). However, even this quantity of assistance would be insufficient to meet the minimum caloric requirements of 2,100 kcal per person per day (49) for the population. This further highlights the critical role of CKs, which serve as the last line of defence against famine for many urban households, especially in hard-to-reach areas.

Even with CK support, our modelling approach indicates that a significant proportion of the population may have consumed less than one meal per day, particularly during periods when kitchens were closed due to violence or funding shortages. Notably the observed drop in kitchens’ funding in November, coincided with a subsequent drop in calories delivered as simultaneously the demand for CKs increased and more beneficiaries attended daily. These same constrains limit coverage of CK interventions in the population served. The population of Jebel Awlia is estimated to be 1.2 million (38), but only 57 CKs are operational in the current constraints, serving a maximum of 233,000 individuals, or 20% of the population. In addition, there was a varying influx of internally displaced people in Jebel Awlia, increasing needs based on very limited resources.

This study demonstrates the feasibility of using invoice-based modelling to monitor food aid distribution and calorie intake in conflict-affected areas, with a particular focus on CKs operating in Jebel Awlia, of Khartoum State. Rather than providing precise calorie intake values, our model estimates ranges and potential trends over time, allowing different food availability scenarios to be explored under different assumptions (Figure 6). This trend-based approach allows to understand how food access may evolve in response to changing resources or disruptions, highlighting periods of potential scarcity. This approach differs from current food aid monitoring mechanisms, which are generally implemented as part of top-down approaches led by UN agencies or international NGOs (50,51). These conventional methods often rely on periodic retrospective assessments, centralised reporting and logistical data from large-scale food distributions, which can be delayed, resource-intensive and disconnected from real-time local dynamics. In contrast, our approach relies on routinely generated field data (52,53)—in particular local food purchase invoices—to produce a more responsive and detailed picture of food aid provision. This basic methodology not only can democratise research and learning for local actors but improves transparency and provides a scalable model that can be adapted in similar contexts where formal monitoring mechanisms are limited or disrupted.

Although based on operational data, another contribution of our study lies in providing a dynamic estimate of calorie intake among vulnerable populations, a measure that remains difficult to achieve in humanitarian contexts. Few tools currently exist to quantify calorie consumption continuously and accurately at local level (54). The Food Consumption Score (FCS) (55), an indicator for food security, offers retrospective and standardized measures of dietary diversity and frequency, but does not provide continuous or direct estimates of caloric intake. Consequently, methods such as invoice-based modelling represent a complementary approach that enables both systematic monitoring of food availability and retrospective estimation of caloric intake from existing operational records.

While this approach provides valuable information on food availability over time, it additionally underscores both the strengths and constraints of locally led food aid interventions in an operationally complex environment. CKs, particularly in contexts such as Jebel Awlia, offer a flexible, rapidly deployable and locally designed mechanism for addressing acute food insecurity, however they have been frequently disrupted by funding shortages, insecurity, and telecommunication blackouts (56–58). Nevertheless, they adapt to changing security conditions, build on local supply chains and can encourage transparency and cohesion in communities by directly involving neighbourhood members in the planning and delivery of services. Yet, this approach is not without its limitations. Firstly, data collected via WhatsApp messages, while efficient and accessible, can be subject to reporting bias, inconsistent formatting and incomplete records. Secondly, data based on invoices often lacks precision in quantities, for example, ‘a bag of rice’ may vary in weight or quality, making it difficult to standardise estimates of food volumes and calories across different kitchens and reliant on seeking standard measures. Thirdly, while invoices provide some indication of the volume of food purchased, they do not always specify the timing of distributions, requiring assumptions about the number of days over which food is consumed or shared. These uncertainties limit the temporal resolution of the analysis and can lead to errors in estimating daily calorie intake per person. For example, these limitations have contributed to the wide confidence intervals observed in the calorie intake estimates for February 2025 (Figure 4). Finally, while we recognize the limitations of this reconstruction, the absence of up-to-date food security monitoring made it necessary to use this method to estimate baseline caloric intake. Although our approach considers only one source of food - community kitchen distributions - in a context of extreme food shortage, we cannot make assumptions about the relative contributions of other sources, such as market purchases (via personal income or remittances) or additional food aid obtained through informal mechanisms such as in-kind support or other social networks.

By partnering directly with kitchen operators and tracking distribution patterns over time, we have been able to piece together a dynamic picture of food access in an area that has remained largely invisible to humanitarian monitoring systems. Our approach not only offers a methodology for estimating caloric intake, but also a practical framework for utilising relatively simple operational data to improving real-time food monitoring in contexts of insecurity and data scarcity. With improved monitoring of food consumptions, user dependency, and more information on kitchen operations, similar models could be adapted to other areas, helping inform local actors and international partners on the impact of community kitchens. Furthermore, enabling more robust learning for planning, adapting, and prioritising response to meet the needs of the population.

## Conclusions

This study proposes a novel and pragmatic method for quantifying food aid delivery and estimating caloric intake through the prism of community kitchen operations. Drawing on invoice records, we were able to provide a unique snapshot of food provision in Jebel Awlia, one of the most food insecure areas of Khartoum State. Our findings confirm that community kitchens are not only essential for maintaining food supplies for vulnerable populations but are often the only remaining safety net in a context of severe household vulnerability and markedly restricted international aid. Yet despite their essential role, their reach remains limited by insecurity, limited resources and inconsistent fiscal support. The siege-like conditions in Jebel Awlia mean that many people may have faced extreme food shortages, consuming far less than the minimum daily caloric requirement. This research highlights the urgent need to scale up support for locally led food aid, not as a stopgap, but as a central pillar of emergency nutrition strategies in Sudan and potentially other complex humanitarian crises. It also highlights the value of collaborative work directly with local actors to capture real-time data, drive accountability, and enable a response that can be planned and adapted based on the ground reality.

## Data Availability

The data and code are available on request at the following github link: https://github.com/yamnao/food_extraction_sudan

## Acknowledgement

We acknowledge the team at Hadhreen for their invaluable contribution to data collection and for providing access to the datasets that were essential to this study. For the purposes of open access, the author has applied a Creative Commons Attribution (CC BY) licence to any Accepted Author Manuscript version arising from this submission.

## References

1. Crisis in Sudan: What is happening and how to help | International Rescue Committee (IRC) [Internet]. 2025 [cited 2025 Apr 13]. Available from: https://www.rescue.org/uk/article/crisis-sudan-what-happening-and-how-help

2. GOV.UK [Internet]. 2023 [cited 2025 Apr 13]. Sudan is now one of the worst countries in the world for humanitarian access: UK statement at the Security Council. Available from: https://www.gov.uk/government/speeches/sudan-is-now-one-of-the-worst-countries-in-the-world-for-humanitarian-access-uk-statement-at-the-security-council

3. Annual Country Report | World Food Programme [Internet]. [cited 2025 Apr 25]. Available from: https://www.wfp.org/operations/annual-country-report?operation_id=SD02&year=2022

4. Ahmed K, Johnson S. Sudan had largest number of people facing extreme food shortages in 2023, UN report shows. The Guardian [Internet]. 2024 Apr 24 [cited 2025 Apr 13]; Available from: https://www.theguardian.com/world/2024/apr/24/sudan-extreme-food-shortages-2023-food-insecurity

5. Sudan faces unprecedented hunger and displacement as war enters third year | UN News [Internet]. 2025 [cited 2025 Apr 13]. Available from: https://news.un.org/en/story/2025/04/1162096

6. Bank AD. African Development Bank Group. African Development Bank Group; 2019 [cited 2025 Mar 25]. Statistics publications. Available from: https://www.afdb.org/en/knowledge/statistics/publications

7. (PDF) Assessment of Food and Nutrition Insecurity using Food Consumption Data in Central Sudan [Internet]. [cited 2025 Apr 25]. Available from: https://www.researchgate.net/publication/366614501_Assessment_of_Food_and_Nutrition_Insecurity_using_Food_Consumption_Data_in_Central_Sudan

8. Sudan Rural Household Survey 2023: Sampling and implementation procedures for the first round [Internet]. [cited 2025 Feb 10]. Available from: https://ebrary.ifpri.org/digital/collection/p15738coll2/id/137139/

9. Global Focus [Internet]. [cited 2025 Apr 13]. Sudan situation. Available from: https://reporting.unhcr.org/operational/situations/sudan-situation

10. Sudan emergency [Internet]. [cited 2025 Apr 13]. Available from: https://www.who.int/emergencies/situations/sudan-emergency

11. Khogali A, Homeida A. Impact of the 2023 armed conflict on Sudan’s healthcare system. Public Health Chall. 2023;2(4):e134.

12. GOV.UK [Internet]. [cited 2025 Apr 13]. Country policy and information note: humanitarian situation, Sudan, February 2024 (accessible). Available from: https://www.gov.uk/government/publications/sudan-country-policy-and-information-notes/country-policy-and-information-note-humanitarian-situation-sudan-february-2024-accessible

13. SMART Methodology [Internet]. [cited 2025 Apr 13]. About SMART. Available from: https://smartmethodology.org/about-smart/

14. Kent L. CNN. 2025 [cited 2025 Apr 25]. A US-run system alerts the world to famines. It’s gone dark after Trump slashed foreign aid. Available from: https://www.cnn.com/2025/03/09/world/us-foreign-aid-freeze-famine-fewsnet-intl/index.html

15. National Public Health Information Coalition (NPHIC) - Impact of FEWS NET Shutdown on Global Famine Response [Internet]. [cited 2025 Apr 25]. Available from: https://www.nphic.org/news/news-highlights/2272-impact-of-fews-net-shutdown-on-global-famine-response?utm_source=chatgpt.com

16. BBC News [Internet]. 2025 [cited 2025 Apr 25]. Sudan conflict: USAID cut hits people ‘screaming from hunger’. Available from: https://www.bbc.com/news/articles/cy7×87ev5jyo

17. ACAPS Thematic Report: Sudan - Humanitarian access developments (October 2024 to March 2025) (10 April 2025) - Sudan | ReliefWeb [Internet]. 2025 [cited 2025 Apr 13]. Available from: https://reliefweb.int/report/sudan/acaps-thematic-report-sudan-humanitarian-access-developments-october-2024-march-2025-10-april-2025

18. Olson S., Dahab M, Parker M. Key Considerations: Mutual Aid Lessons and Experiences From Emergency Response Rooms in Sudan [Internet]. The Institute of Development Studies and Partner Organisations; 2024 Oct [cited 2025 Apr 25]. Available from: https://opendocs.ids.ac.uk/articles/report/Key_Considerations_Mutual_Aid_Lessons_and_Experiences_From_Emergency_Response_Rooms_in_Sudan/27292296/1

19. Sudan: Community kitchens bring vital food relief to thousands in North Darfur as humanitarian conditions worsen[EN/AR] - Sudan | ReliefWeb [Internet]. 2024 [cited 2025 Apr 25]. Available from: https://reliefweb.int/report/sudan/sudan-community-kitchens-bring-vital-food-relief-thousands-north-darfur-humanitarian-conditions-worsenenar

20. The New Humanitarian | ‘We survive together’: The communal kitchens fighting famine in Khartoum [Internet]. 2024 [cited 2025 Apr 25]. Available from: https://www.thenewhumanitarian.org/news-feature/2024/06/24/we-survive-together-communal-kitchens-fighting-famine-khartoum-sudan

21. Hadhreen [Internet]. [cited 2025 May 15]. Available from: https://hadhreen.org/ar/

22. https://hadhreen.org/wp-content/uploads/sites/3/2025/05/Hadhreen-2022-annual-report.pdf [Internet]. [cited 2025 May 15]. Available from: https://hadhreen.org/wp-content/uploads/sites/3/2025/05/Hadhreen-2022-annual-report.pdf

23. Hadhreen-2023-annual-report.pdf [Internet]. [cited 2025 May 15]. Available from: https://hadhreen.org/test2025/wp-content/uploads/sites/3/2025/05/Hadhreen-2023-annual-report.pdf

24. SudanTribune. Sudan Tribune. 2025 [cited 2025 Apr 29]. Sudanese army secures strategic Jebel Aulia dam. Available from: https://sudantribune.com/article299152/

25. Sudan conflict: warring sides blame each other for strike on key bridge | Sudan | The Guardian [Internet]. [cited 2025 Jun 3]. Available from: https://www.theguardian.com/world/2023/nov/18/sudan-conflict-strike-damages-dam-bridge-in-latest-key-infrastructure-damage

26. Alleged drone strike hits Jebel Aulia market [Internet]. Centre for Information Resilience. 2024 [cited 2025 Jun 3]. Available from: https://www.info-res.org/sudan-witness/reports/flash-report-alleged-drone-strike-hits-jebel-aulia-market/

27. Food and nutrition crisis deepens across Sudan as famine identified in additional areas [Internet]. [cited 2025 Feb 13]. Available from: https://www.unicef.org/press-releases/food-and-nutrition-crisis-deepens-across-sudan-famine-identified-additional-areas

28. https://www.ipcinfo.org/fileadmin/user_upload/ipcinfo/docs/IPC_Sudan_Acute_Food_Insecurity_Oct2024_May2025_Snapshot.pdf [Internet]. [cited 2025 Apr 28]. Available from: https://www.ipcinfo.org/fileadmin/user_upload/ipcinfo/docs/IPC_Sudan_Acute_Food_Insecurity_Oct2024_May2025_Snapshot.pdf

29. Highlight 11 August 2023 | United Nations Secretary-General [Internet]. [cited 2025 Jun 3]. Available from: https://www.un.org/sg/en/content/highlight/2023-08-11.html

30. Daily Press Briefing by the Office of the Spokesperson for the Secretary-General, 27 December 2024 - Sudan - Sudan | ReliefWeb [Internet]. 2024 [cited 2025 Feb 13]. Available from: https://reliefweb.int/report/sudan/daily-press-briefing-office-spokesperson-secretary-general-27-december-2024-sudan

31. UNDP [Internet]. [cited 2025 Feb 13]. Rapid Assessment: Economic Situation of Urban Population in Khartoum State. Available from: https://www.undp.org/sudan/publications/rapid-assessment-economic-situation-urban-population-khartoum-state

32. ISO [Internet]. [cited 2025 May 6]. ISO - ISO 3166 — Country Codes. Available from: https://www.iso.org/iso-3166-country-codes.html

33. Sudan | OCHA [Internet]. 2025 [cited 2025 May 15]. Available from: https://www.unocha.org/sudan

34. MarianMT — transformers 3.5.0 documentation [Internet]. [cited 2025 May 15]. Available from: https://huggingface.co/transformers/v3.5.1/model_doc/marian.html

35. NutVal (Version 4) | ENN [Internet]. [cited 2025 Apr 25]. Available from: https://www.ennonline.net/projects/nutval-version-4

36. publisher E. World Health Organization - Regional Office for the Eastern Mediterranean. [cited 2025 Apr 28]. Sudan launches comprehensive food composition tables. Available from: http://www.emro.who.int/sdn/sudan-news/sudan-launches-comprehensive-food-composition-tables.html

37. Moahmed TA, El Gayar N, Atiya AF. Forward and Backward Forecasting Ensembles for the Estimation of Time Series Missing Data. In: El Gayar N, Schwenker F, Suen C, editors. Artificial Neural Networks in Pattern Recognition. Cham: Springer International Publishing; 2014. p. 93–104.

38. World Population Dashboard -Sudan | United Nations Population Fund [Internet]. [cited 2025 Feb 10]. Available from: https://www.unfpa.org/data/world-population/SD

39. Raleigh C, Linke A, Hegre H, Karlsen J. Introducing ACLED: An Armed Conflict Location and Event Dataset. J Peace Res. 2010;47(5):651–60.

40. R: a language and environment for statistical computing [Internet]. [cited 2025 Feb 24]. Available from: https://www.gbif.org/tool/81287/r-a-language-and-environment-for-statistical-computing

41. The Python Language Reference — Python 2.7.18 documentation [Internet]. [cited 2025 Apr 25]. Available from: https://docs.python.org/2/reference/

42. Vitamin and mineral requirements in human nutrition, 2nd edition [Internet]. [cited 2025 Apr 25]. Available from: https://www.who.int/publications/i/item/9241546123

43. Al-kalakla Emergency Room’s Post [Internet]. 2023. Available from: https://www.facebook.com/permalink.php?story_fbid=pfbid08HDM2rhmgznfMY4W3NZU8raWpD SsjEwtRt4izNbyeZWC6N8AuqBfFQUZc1Sq6YvEl&id=100092756166266

44. Facebook Post [Internet]. 2024 .?????? ???? ????? ????. Available from: https://www.facebook.com/photo/?fbid=283855948016976&set=pb.100091777192729.-2207520000

45. Highlight 11 August 2023 | United Nations Secretary-General [Internet]. [cited 2025 Apr 29]. Available from: https://www.un.org/sg/en/content/highlight/2023-08-11.html

46. Sudan: Humanitarian Access Snapshot (December 2024) - Sudan | ReliefWeb [Internet]. 2025 [cited 2025 Apr 29]. Available from: https://reliefweb.int/report/sudan/sudan-humanitarian-access-snapshot-december-2024

47. Sudan: Humanitarian Access Snapshot (December 2024) - Sudan | ReliefWeb [Internet]. 2025 [cited 2025 Feb 13]. Available from: https://reliefweb.int/report/sudan/sudan-humanitarian-access-snapshot-december-2024

48. Operational Update on surge in food aid in Sudan | World Food Programme [Internet]. 2024 [cited 2025 Feb 10]. Available from: https://www.wfp.org/news/operational-update-surge-food-aid-sudan

49. Griekspoor A, Collins S. Raising standards in emergency relief: how useful are Sphere minimum standards for humanitarian assistance? BMJ. 2001 Sep 29;323(7315):740–2.

50. Paulus D, de Vries G, Janssen M, Van de Walle B. Reinforcing data bias in crisis information management: The case of the Yemen humanitarian response. Int J Inf Manag. 2023 Oct 1;72:102663.

51. Keynote Paper: Individual food intake survey methods [Internet]. [cited 2025 Apr 29]. Available from: https://www.fao.org/4/y4249e/y4249e0a.htm

52. Time to Listen: Hearing People on the Receiving End of International Aid - CDA Collaborative Learning [Internet]. [cited 2025 Apr 29]. Available from: https://www.cdacollaborative.org/publication/time-to-listen-hearing-people-on-the-receiving-end-of-international-aid/

53. ALNAP [Internet]. 2016 [cited 2025 Apr 29]. Listening to Communities in Insecure Environments - Lessons from Community Feedback Sessions in Afghanistan, Somalia & Syria. Available from: https://alnap.org/help-library/resources/listening-to-communities-in-insecure-environments-lessons-from-community-feedback/

54. How do Different Indicators of Household Food Security Compare? Empirical Evidence from Tigray - World | ReliefWeb [Internet]. 2013 [cited 2025 Jun 2]. Available from: https://reliefweb.int/report/world/how-do-different-indicators-household-food-security-compare-empirical-evidence-tigray

55. ResearchGate [Internet]. [cited 2025 Jun 2]. (PDF) Validation of the World Food Programme’s food consumption score and alternative indicators of household food security. Available from: https://www.researchgate.net/publication/46442105_Validation_of_the_World_Food_Programme’s_food_consumption_score_and_alternative_indicators_of_household_food_security

56. The New Humanitarian | Communication blackout thwarts mutual aid efforts in besieged Khartoum [Internet]. 2024 [cited 2025 Apr 29]. Available from: https://www.thenewhumanitarian.org/news-feature/2024/03/04/sudan-communication-blackout-mutual-aid-efforts-besieged

57. The New Humanitarian | Sudan mutual aid groups face survival battle amid army abuse and US aid freeze [Internet]. 2025 [cited 2025 Apr 29]. Available from: https://www.thenewhumanitarian.org/news-feature/2025/02/26/sudan-mutual-aid-groups-face-survival-battle-amid-army-abuse-and-us-aid

58. Information Saves Lives | Internews [Internet]. [cited 2025 Apr 29]. Information Crisis Response in Sudan and South Sudan. Available from: https://internews.org/areas-of-expertise/humanitarian/projects/conflict-projects/information-crisis-response-in-sudan-and-south-sudan/

